# A Graph Embedding Approach for Deciphering the Longitudinal Associations of Global Mobility and COVID-19 Cases

**DOI:** 10.1101/2023.03.30.23287969

**Authors:** Raghav Awasthi, Meet Modi, Hardik Dudeja, Tanav Bajaj, Shruti Rastogi, Tavpritesh Sethi

## Abstract

The COVID-19 pandemic has highlighted the importance of monitoring mobility patterns and their impact on disease spread. This paper presents a methodology for developing effective pandemic surveillance systems by extracting scalable graph features from mobility networks. We utilized Meta’s “Travel Patterns” dataset to capture the daily number of individuals traveling between countries from March 2020 to April 2022. We have used an optimized node2vec algorithm to extract scalable features from the mobility networks. Our analysis revealed that movement embeddings accurately represented the movement patterns of countries, with geographically proximate countries exhibiting similar movement patterns. The temporal association dynamics between Global mobility and COVID-19 cases highlighted the significance of high-page rank centrality countries in mobility networks as a key intervention target in controlling infection spread. Our proposed methodology provides a useful approach for tracking the trajectory of infectious diseases and developing evidence-based interventions.

## Introduction

Pandemic outbreaks pose devastating consequences on public health, economies, and social structures [1]. Early detection and rapid response are crucial for mitigating their impact, which requires the timely identification of infected individuals, effective contact tracing, and isolation measures. However, traditional surveillance methods, such as laboratory testing and manual contact tracing, can be slow, labor-intensive, and limited in their scope [2]. With the advancement of technology, data-driven approaches have become increasingly crucial for disease surveillance. In particular, mobility networks, which capture human movement patterns, can provide valuable information for pandemic surveillance [3]. Mobility networks can be constructed from various data sources, such as mobile phone location data, transportation logs, and social media check-ins. Mobility networks are essential for pandemic surveillance. They can provide valuable information about how diseases are spreading geographically and help public health officials make informed decisions about allocating resources and implementing mitigation measures. By analyzing mobility networks, researchers can gain insights into the spread of infectious diseases, identify high-risk areas, and inform public health interventions. However, analyzing and extracting useful information from mobility networks can be challenging due to their complex structure. Mobility networks can be represented as graphs, where nodes represent locations or individuals, and edges represent movement or connections between them. Graphs can have millions or even billions of nodes and edges, making traditional analysis methods impractical.

There has been significant research on using mobility networks for pandemic surveillance [4] [5]. One of the earliest examples of this approach [6], used mobile phone data to construct a network of human mobility and simulated the spread of influenza in the United States. They found that mobility networks can provide valuable insights into the spread of infectious diseases and that targeted interventions based on mobility patterns can be more effective than random interventions. Since then, numerous studies have used mobility networks for pandemic surveillance, focusing on different aspects of the problem. For example, researchers used mobile phone data to construct a mobility network that can predict the spread of malaria [7] [3]. However, analyzing and extracting useful information from mobility networks can be challenging due to their large scale and complexity. To overcome this challenge, researchers have proposed using scalable graph features to summarize the information contained in mobility networks. In this research, we propose to learn scalable graph features from mobility networks to construct robust pandemic surveillance. The objective is to develop an efficient approach that can extract relevant information from mobility networks in a scalable and effective manner. By doing so, we hope to contribute to developing effective pandemic surveillance systems that can aid in the early detection and control of infectious disease outbreaks.

## Method

This paper proposes a node embedding methodology (**Figure 1**) to extract scalable features from mobility networks using COVID-19 as a use case. We also presented various methods to evaluate the effectiveness of the extracted features by analyzing their correlation with attributes relevant to modeling COVID-19 pandemic policies. Our approach is suitable for tracking the trajectory of infection in other infectious diseases and provides a strong basis for evidence-based interventions.

**Figure 1:**
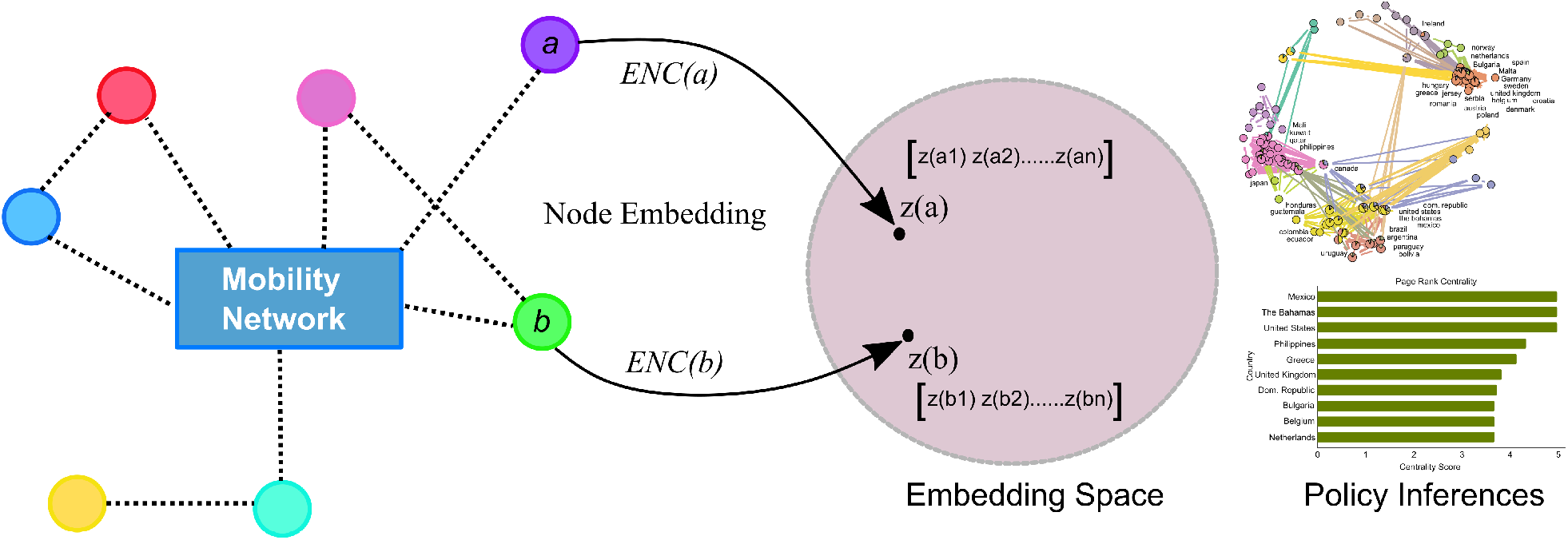
**Pipeline depicts our methodology for analyzing mobility network data**; we used the COVID-19 country-level mobility pandemic as an example. We used the node2vec method to extract the distinctive features of each country. By integrating these features with country-specific COVID-19 indicators, we identified able to determine policy implications, such as the need for travel restrictions.

### Node Embedding

Node embeddings are vector representations of nodes in a network or graph. Mathematically, we defined node embedding for a graph *G* on *N* nodes with vertex set *V* (*G*) = {*v*1, *v*2, …, *vN*} as d-dimensional (generally *d < N*) representations of the nodes [8] [9]. These embeddings capture the topology of graphs and are frequently utilized as features of machine learning algorithms to model outcomes. Node embedding algorithms are a mapping function *f* : *V* (*G*) → *R*^*d*^. To learn *f*, a neighborhood sampling strategy *S* is used, which generates a network neighborhood of node *u* defined as *NS*(*u*) ⊂ *V* for every source node *u* ∈ *V*. The objective function mentioned below is optimized by the algorithm, which maximizes the log-probability of observing a network neighborhood *NS*(*u*) for a node conditioned on its feature representation,

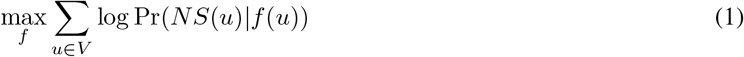

The embedding algorithms make the following two assumptions:

#### Conditional independence

This states that the observation of any other node does not influence the chance of observing a node in a neighborhood.

#### Symmetry in feature space

This states that there is symmetry between the source node and the neighborhood node, meaning that they have equal and opposite effects on each other.

These assumptions simplify the above equation to:

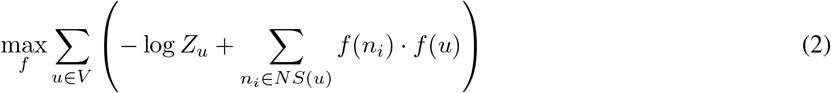

The per-node partition function is and [10] [11]

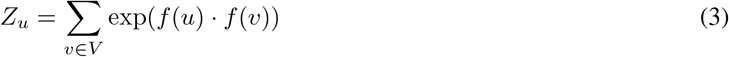

### Classical Search Strategy

The optimization of **Eqaution 2** involves the selection of neighborhood nodes surrounding a source node through neighborhood search strategies. The algorithms defined below are popular algorithms that suggest a solution to limit the neighborhood size and generate multiple sample neighborhood sets for a given node, *u*.

#### Breadth First Search (BFS)

This approach confines the neighborhood to the source’s immediate neighbors.

#### Depth First Search (DFS)

This approach includes nodes at progressively greater distances from the source node.

### Node2Vec

node2vec [12] is a node embedding algorithm that learns low-dimensional representations of nodes in a network by optimizing an objective function that captures the structural properties of the graph. node2vec builds upon the Breadth-

First Search (BFS) and Depth-First Search (DFS) algorithms. Specifically, it uses a biased random walk strategy to generate node sequences that capture the graph’s local and global structural information. During the random walk, the node2vec algorithm chooses the next node to visit based on a probability distribution that balances BFS-like behaviour and DFS-like behaviour. This allows node2vec to capture both the local connectivity patterns of nodes (BFS-like behaviour) and the long-range structural dependencies between nodes (DFS-like behaviour). After generating the node sequences using the random walk strategy, node2vec applies a skip-gram model to learn low-dimensional representations of nodes optimized to predict each node’s context (i.e. the nodes that appear in its neighborhood).

### Policy Inferences from Mobility Networks Utilizing Node Emdedding

Understanding mobility networks is challenging due to their complex patterns of movement and connectivity between nodes, which can be influenced by multiple factors such as travel preferences, socioeconomic status, and infrastructure. These networks consist of nodes representing geographic locations and edges representing movement links. node2vec can address this challenge by learning low-dimensional representations of nodes in a network. Below is a list of possible policy inferences that can be effectively derived from mobility network embedding.

1. **Target interventions to high-risk areas** Policymakers can use data on mobility patterns to identify high-risk areas and develop targeted interventions to reduce the spread of the virus. This can include increased cleaning and sanitation measures or restrictions on the number of people allowed in certain areas.
2. **Resource allocation policies** Mobility networks and node embedding may be used to create more efficient resource allocation strategies. This strategy may aid in the identification of high-traffic locations, the response to dynamic situations, and the assistance of vulnerable groups.
3. **Lockdown and Travel Restriction policies** Node embedding and mobility network analysis may help formulate appropriate pandemic lockdown and travel restriction policies. It can identify high-risk areas, evaluate the impact of restrictions on mobility, and inform targeted interventions.
4. **Effective vaccination policy** Policymakers can maximise vaccination distribution and outreach by assessing movement patterns and high-risk locations.

## Results

The results section of this paper presents the use of the node2vec method on mobility networks in the context of COVID-19. The experiments began with optimizing node2vec model hyperparameters, followed by analyzing the spatiotemporal patterns and community learning from the learned node embedding features within the context of COVID-19.

### Data

Our study utilized the “Travel Patterns” dataset provided by Facebook (now Meta). This dataset records the daily number of Facebook users who travel between countries, and it was collected from individuals who voluntarily shared their location data with the Facebook app. The dataset includes those who have travelled over long distances, such as by air or train. The Travel Patterns dataset is especially useful for epidemiological study, as it can aid in understanding the impact of international travel on disease spread as well as provide insights into the economic consequences of reduced travel during public health emergencies and other events. We analyzed data from March 2020 to April 2022 to cover all the major waves of COVID-19, including alpha, beta, gamma, delta, and omicron. Also, we have used country-wise daily COVID-19 data, which we have extracted from a publicly available data repository *World Health Organization (WHO)*. ^1^

### Data Preprocessing

The country-level longitudinal data downloaded from the *Meta* portal and *WHO* website were originally tabular, and we performed the following preprocessing steps to prepare the data for analysis.

#### Mobility Data

Considering the commonly held theory that COVID-19-infected individuals usually experience symptoms, such as mild respiratory symptoms and fever, within 1–14 days [13], we analyzed movement between countries on a biweekly basis. We summarized the country-to-country movement biweekly by taking the mean of the daily movement.

#### Covid-19 data

We have also summarized the COVID-19 daily new cases on a biweekly basis.

### Hyperparameter-optimized node2vec learning

Optimizing the node2vec algorithm involves adjusting several hyperparameters, among which the embedding dimension is one of the most significant. The embedding dimension determines the number of features in the node vector representation. Optimizing it can enhance the quality of embeddings and the overall performance of subsequent tasks [14] [15]. This study aimed to determine the optimal embedding dimension for the node2vec algorithm in the context of a bi-weekly mobility network and COVID-19 case prediction. To achieve this, we embedded the mobility network into dimensions ranging from 5 to 50, with a 5-unit increment, where each node represented a country. The maximum value of 50 was selected based on the size of the smallest network in our mobility network, which had 54 nodes. Using the embedded features, we then integrated the next 15 days’ COVID-19 cases and employed a regression approach to train machine learning models (random forest and xgboost) [16] [17]. We divided the dataset into training and testing sets at an 80:20 ratio and then trained the model using five-fold cross-validation on the training set. We evaluated the model performance score (R2) on the testing set. 95% confidence interval of R2 socre was calculated using **Equation 4**. Our findings revealed that for both random forest and xgboost models, the R2 increased with the embedding dimension and then started decreasing (**Figure 2A, Figure 2B**). We identified the highest R2 at embedding dimensions 25 for random forest and xgboost. Therefore, we selected the optimal embedding dimension of 25 to preserve the maximum information. Additionally, we calculated the top 5 features (**Figure 2C, Figure 2D**) from the random forest and xgboost models with the optimum embedding dimension of 25. We found that both models had 80% of their top features in common, indicating consistency in the learned machine-learning models.

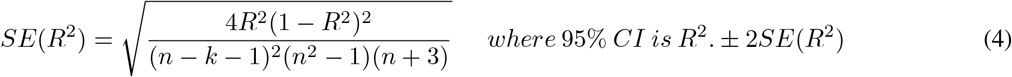

**Figure 2:**
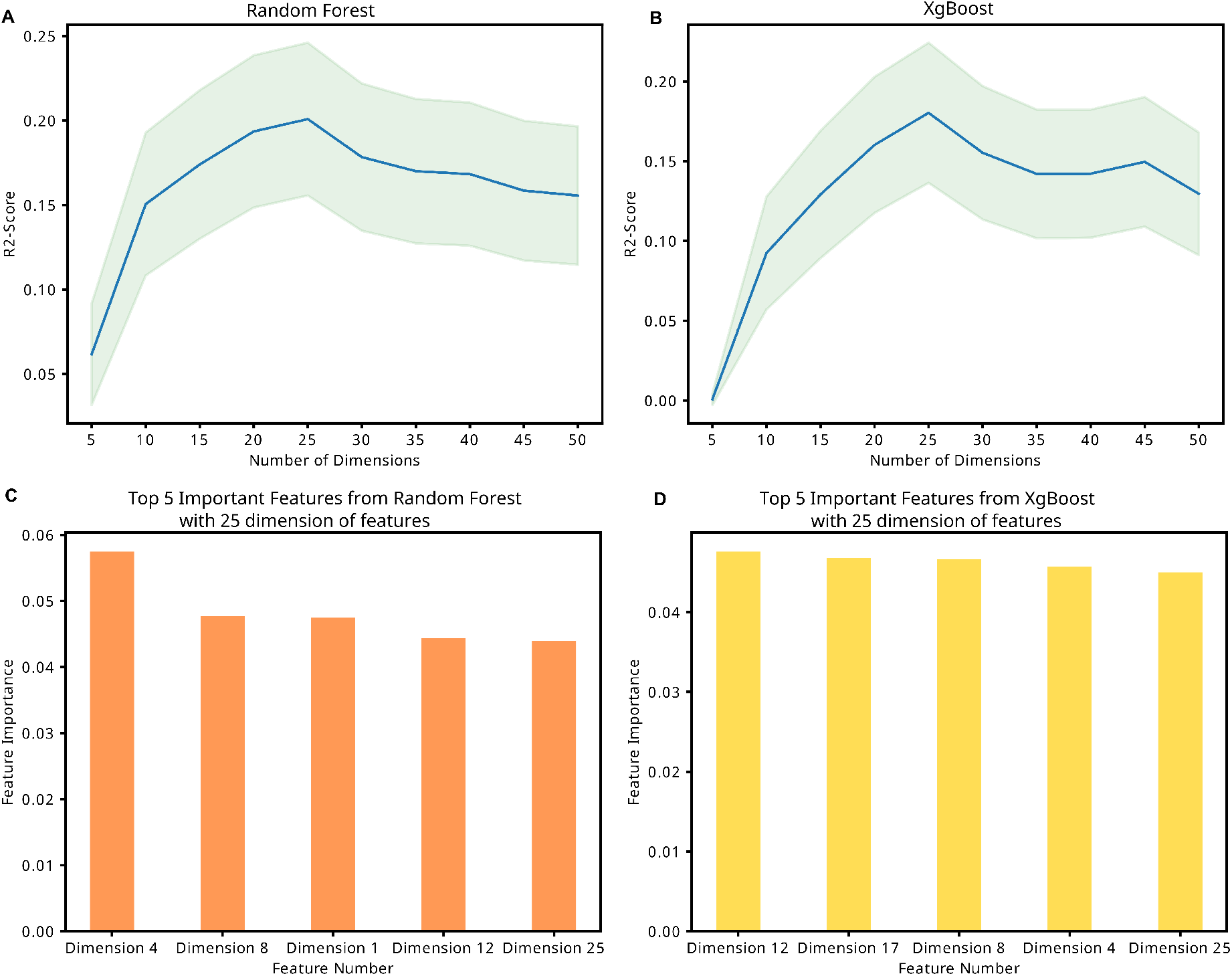
Node2Vec Learning. (A) R2 score with 95% CI for the prediction of the next 15 days Cases of COVID-19 from the Random Forest model with an embedding dimension of 5 to 50 (B) R2 score for predicting the next 15 days of COVID-19 cases from the XGBoost model with an embedding size of 5 to 50 C) The top five features of a Random Forest model with the optimal embedding dimension D) The top five features of n XGBoost model with the optimal embedding dimension

### Discovering Spatiotemporal Patterns in Embedded Features: A Way to Enhance Travel Restrictions and Resource Allocation Policies

Learning the effective implementation of policies like travel restrictions, resource allocation, and social distancing during a pandemic is a challenging task due to the unpredictable nature of the pandemic, the emergence of new variants, and the complex interactions between socioeconomic and geopolitical factors related to the pandemic. But analyzing spatiotemporal mobility patterns could help policymakers make better decisions. However, the asymmetric proximity scores (mobility from country A to country B is not the same as mobility from country B to country A) make it hard to analyze the spatiotemporal patterns of the original mobility network. We used the embedded features to calculate the Pearson correlation between countries and built correlation networks to solve this problem.

The temporal dynamics of the correlation networks were learned using an alluvial diagram (**Figure 3A**) and biweekly Jaccard Index calculations between the top 20 countries in the movement network and those with the highest COVID-19 cases (**Figure 3B**). Results showed that the movement embeddings were a meaningful representation of the movement patterns of countries, with countries nearby having similar movement patterns. As COVID-19 threats declined and lockdown policies became lenient, more clusters were formed, indicating increased movement patterns. The Jaccard score also revealed that countries with high page rank centrality were key nodes in the movement embedding correlation network and had higher COVID-19 cases during peaks in the pandemic suggesting that movement patterns play a significant role in determining how the pandemic spreads from one region to another. Also, after a sudden drop in Jaccard Score, there is a gradual increase in its value as soon as government policies change indicating how knowing these trends might help the policymakers in planning for such pandemics effectively.

**Figure 3:**
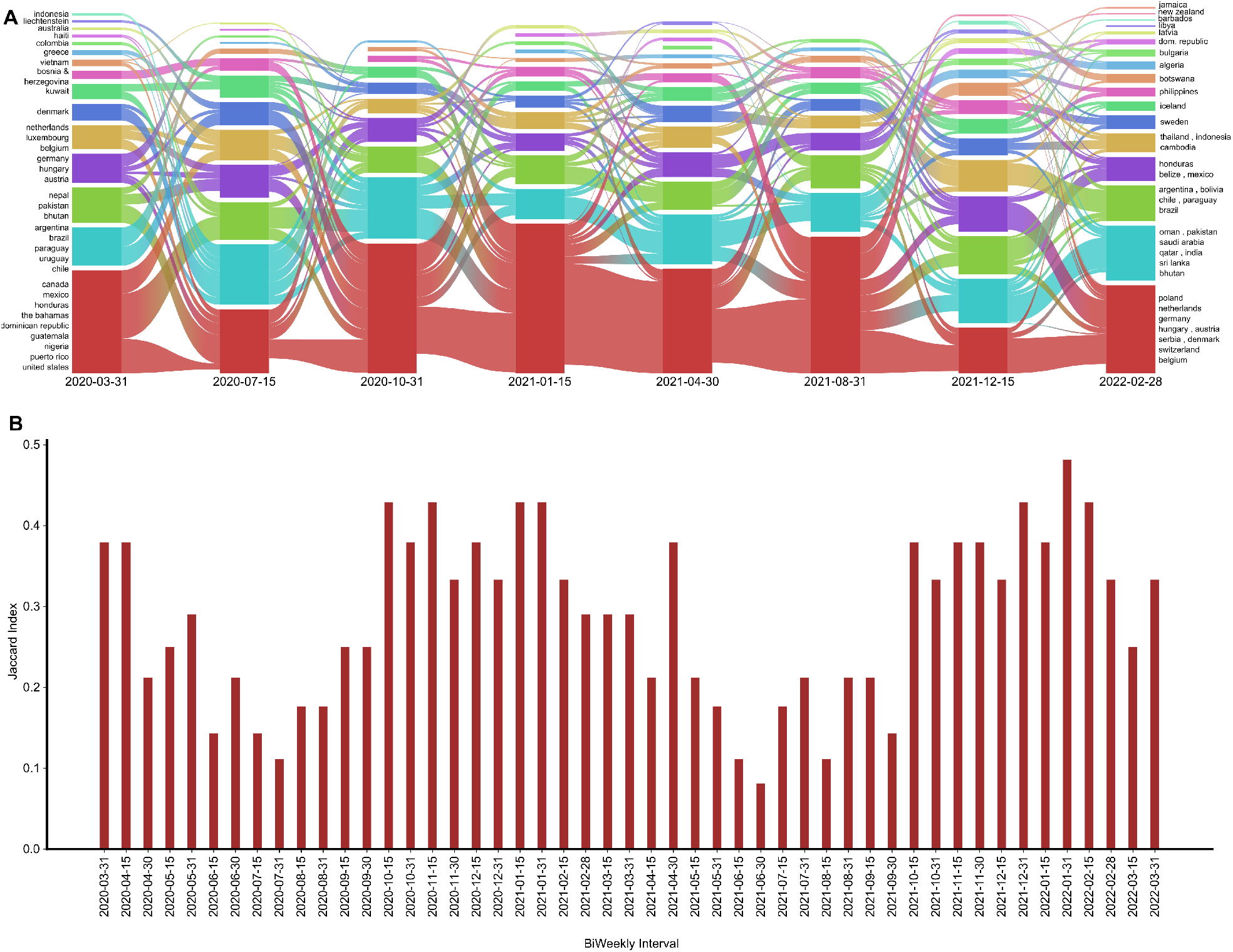
Spatio-temporal representation of the mobility network pattern extracted from the embedding features. A) Alluvial diagram representing the cluster of countries based on movement patterns and their flow from March 2020 to April 2022. B) A biweekly jaccard score between the top 20 countries that were central in the movement network and countries with the 20 highest COVID-19 cases

### Enhancing and Tracking Multilevel Implementation of Interventions through Community Learning in Movement Graphs

Next, We conducted community detection in the mobility network to enhance and track the multilevel implementation of interventions. This can aid in understanding the hierarchical spread of infection and inform strategies to mitigate its impact. [18] [19]. We used a country-wise correlation network learned from an embedded feature and summarized it to create an overall network. To identify communities, we performed a link community algorithm; link communities can unveil the complex structure of networks that is both nested and overlapping, and identify the nodes that play a critical role in connecting different communities.[20] [21]. To perform the analysis, we used R based *linkcomm* [22] package and utilized the McQuitty algorithm [23], which first calculates pairwise distances between all pairs of links in the network (**Equation 5**). The distance between two links is typically defined as the inverse of their similarity.

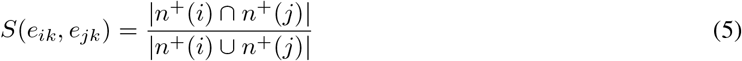

where *e*_*ik*_ and *e*_*jk*_ are link that share a node and *n*^+^(*i*) refers to the first-order node neighbourhood of node *i*,

We found 15 unique communities in the entire movement network using the, with the largest community having 34 nodes. Our findings demonstrate the importance of geographic proximity in COVID-19 mobility patterns by showing that countries that are close by were more likely to belong to the same community. (**Figure 4A, Figure 4B, Figure 4C**). We captured the relationships between overlapping groups and produced a more accurate representation of the COVID-19 movement network by characterizing communities as groupings of links. Using this method, we were able to pinpoint groups of countries that are more connected to one another than to countries in other groups, revealing information about the relationships between countries based on their COVID-19 movement patterns. For instance, European nations, including Germany, the United Kingdom, Sweden, Belgium, and Poland, were seen to be part of the same community. This is also the same for South American nations such as Brazil, Bolivia, Argentina, and Paraguay being in the same community. (Figure 4A) Also, we determined the page rank centrality of the entire movement network (**Figure 4D**), and the results show that Mexico, The Bahamas, and the United States have the highest page rank centralities, followed by the Philippines, Greece, and the United Kingdom. This information can help us comprehend the countries that have contributed significantly to the COVID-19 pandemic spread. This shows essential insights into the COVID-19 movement network’s dynamics and helped identify critical countries for targeted interventions to control the disease’s spread. Overall, this result offers insightful information about the movement and propagation patterns of COVID-19 throughout the pandemic, which can be utilized to guide public health policies and interventions to restrict the disease’s spread.

**Figure 4:**
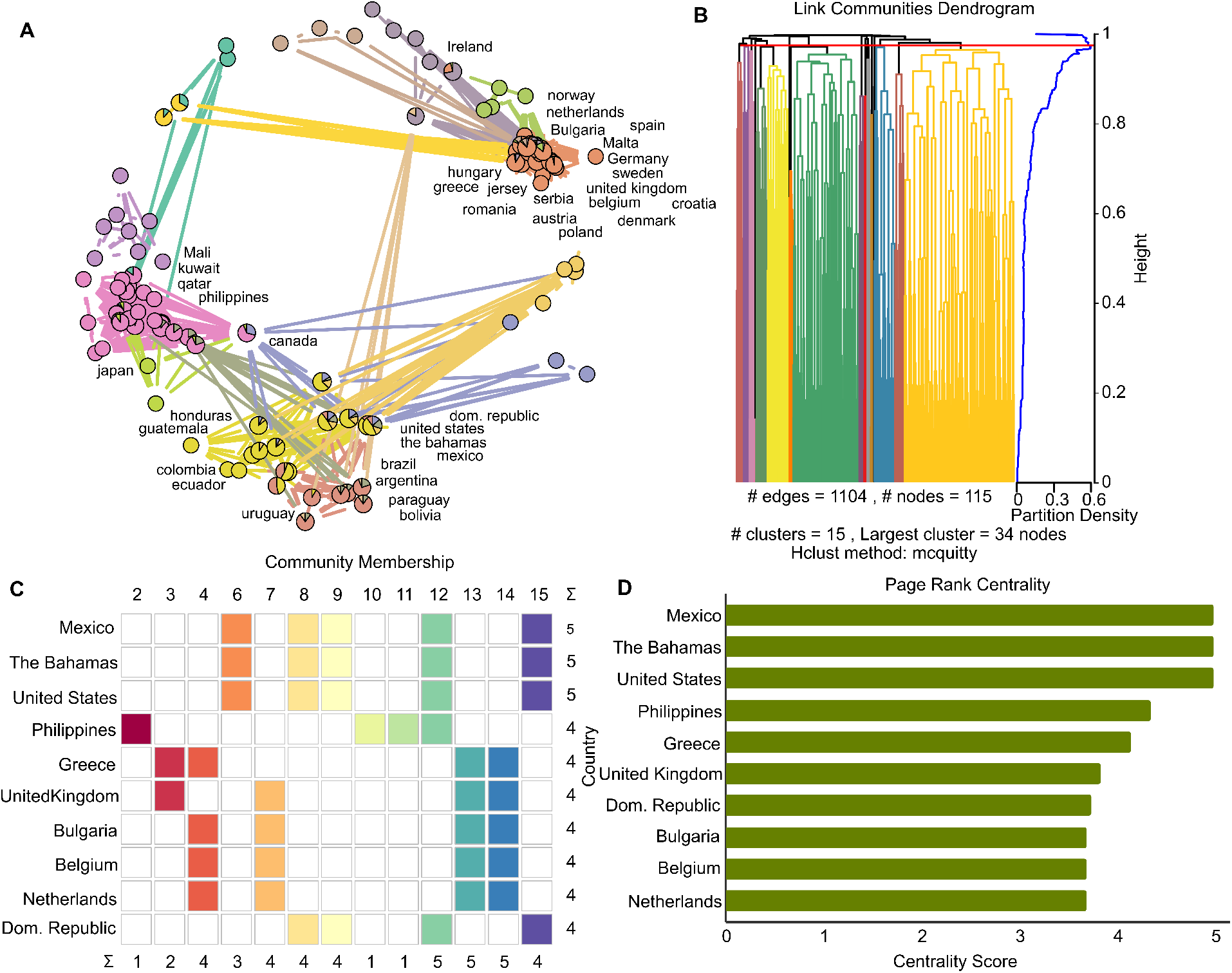
Community learning from the correlation network of embedded features. A) summarized movement network with detected community B) hierarchical clustering in the summarized network to detect communities Community membership of countries with communities representing countries D) Page rank centrality of countries in the overall summarized network

## Conclusion

Graph-based methods have the potential to facilitate infectious disease surveillance more efficiently by strongly comprehending the complexity of data. However, these methods have not been extensively explored or implemented in traditional disease surveillance systems, which rely more on individual-level data, such as symptom reporting or laboratory results. This limitation could be due to the challenges of collecting and processing large-scale network data, a lack of expertise in network science, or limited resources for implementing such approaches in real-time disease surveillance. In this study, we have provided a graph-based algorithm that can understand mobility more extensively and extract scalable features that can be further used for downstream tasks like building lockdown policies, travel restriction policies, forecasting infectious disease trajectories, and vaccination policies. Also, a case study of COVID-19 mobility at the country level showed that the embedded features could predict COVID-19 cases with a good R2 value, which suggests another way to predict how infectious diseases spread. Also, in other results, we’ve detected the groups of countries that show a similar movement. Countries that are close together were found to be in the same group, which shows again how stable embedded features are. From the temporal trend of the Jaccard index, we found that in the time range where the COVID waves were at a peak, the overlap between countries with high movement centrality and countries with high COVID-19 cases is low compared to when COVID-19 cases were not at a peak, which represents that embedded features have nicely captured the travel restriction policies. We also acknowledge that our analysis has some limitations. For example, we used Facebook user data, but Facebook is banned in multiple countries like China, so there might be some missing points in the spatiotemporal analysis. Overall, as per our knowledge, this study is the first to explore the node embedding algorithm for mobility networks. We strongly believe this algorithm can offer a valuable opportunity for enhancing data-driven infectious disease surveillance.

## Data Availability

Travel Patterns data are available to nonprofits and researchers who sign data-sharing agreements with Meta.

https://dataforgood.facebook.com/dfg/tools/travel-patterns#accessdata

## Acknowledgments

We acknowledge support from Facebook (Meta), Delhi Cluster-Delhi Research Implementation and Innovation (DRIIV) Project supported by the Principal Scientific Advisor Office, Prn.SA/Delhi/Hub/2018(C) and the Center of Excellence in Healthcare supported by Delhi Knowledge Development Foundation (DKDF), Prn. SRP206 at IIIT-Delhi. We also acknowledge Abdal Lalit, Onkar Mahapatra, and Urvi Midha for helping us in data collection and data preprocessing.

## Footnotes

1 https://covid19.who.int/data

